# QT prolongation risk factors and a monitoring strategy in rifampicin-resistant tuberculosis: Findings from the STREAM Stage 2 trial

**DOI:** 10.1101/2025.08.08.25333290

**Authors:** G Hughes, E Frangou, H Bern, M Gurumurthy, B Tsogt, CY Chiang, A. J. Nunn, S.K. Meredith, R. L. Goodall, the STREAM Trial Collaborators

## Abstract

**Background:** Treatment of rifampicin-resistant tuberculosis (RR-TB) involves drugs that can prolong the QT interval. There is limited data on risk factors and the level of cardiac monitoring required. We analysed STREAM Stage 2 data to address these issues.

**Methods:** A post-hoc analysis was undertaken of data from participants allocated to 3 regimens: 9-month control (n=202), 9-month oral (n=211) and 6-month (n=143). Risk factors for development of a QT or QTcF interval ≥500ms were assessed. The diagnostic accuracy of a monitoring strategy for QT/QTcF prolongation was tested.

**Results:** QT/QTcF ≥500ms occurred on all regimens: 9-month control (n=14 (6.9%)), 9-month oral (n=8 (3.8%)) and 6-month (n=6 (4.2%)).

The participating country with the highest number of QT/QTcF interval ≥500ms events was Mongolia (18 events [64%]). A higher baseline QTcF was significantly associated with development of QT/QTcF ≥500ms (OR 1.05; 95% CI 1.03 to 1.07, p<0.001). There was a suggested association between moxifloxacin (compared to levofloxacin, OR 2.49; 95% CI 0.92 to 6.71, p = 0.07) and a higher baseline TSH (OR 3.52; 95% CI 0.84 to 14.73, p=0.08) and increased odds of QT/QTcF ≥500ms.

The monitoring strategy performed well in the control (sensitivity 100%; specificity 62%; positive predictive value (PPV) 13% and negative predictive value (NPV) 100%) and oral (sensitivity 100%, specificity 59%, PPV 6% and NPV 100%) regimen groups.

**Conclusions:** Baseline QTcF and country were associated with increased odds of QT/QTcF prolongation. The monitoring strategy performed well in the identification of participants at higher risk of QT/QTcF prolongation.

## Introduction

Cardiac safety remains a key concern in the treatment of drug-resistant tuberculosis (DR-TB). QT or QTcF interval prolongation ≥500ms, which can lead to life-threatening arrythmias, has been associated with several drugs recommended in Groups A and B of the World Health Organisation (WHO) guideline for the management of DR-TB. [1]. In the STREAM Stage 1 trial, 11% (31/282) of participants allocated the study regimen (which included high dose moxifloxacin and clofazimine) developed a QT/QTcF ≥500ms. [2,3]. Analysis of baseline risk factors found a significant association between development of a QT/QTcF ≥500ms and both country (Mongolia) and raised baseline QTcF (≥400ms). [3]. It is unclear whether these factors remain important in a population from a wider range of countries taking the same regimen, or other regimens not included in Stage 1.

Due to cardiac safety concerns in patients taking DR-TB treatment, ECG monitoring remains a key part of clinical management, but there is currently no universally agreed ECG monitoring strategy. This is highlighted by several recent trials. [4–6]. Analysis of STREAM Stage 1 data found an ECG monitoring strategy using a combination of QTcF values of ≥425ms at 4 hours after the first dose and ≥430ms at week 3, successfully identified most participants who reached 500ms with a sensitivity of 97% whilst allowing a reduction in the frequency of ECG monitoring visits for two-thirds of patients who did not reach 500ms. [7] This work raised the possibility of targeted monitoring to identify “high-risk” patients and reduced monitoring on those deemed “lower-risk” based on QT/QTcF values within one month of treatment, which requires validation in a different data set.

The STREAM Stage 2 trial was a phase 3 randomised controlled trial across 13 sites in seven countries. The primary comparison was between a 9-month oral regimen (including bedaquiline, levofloxacin and clofazimine) and the 9-month STREAM Stage 1 regimen (including moxifloxacin or levofloxacin, and clofazimine) which was the control. Assessment of a six-month regimen was a key secondary objective. [8] In Stage 2, levofloxacin replaced moxifloxacin in the control arm in later versions of the protocol ( ≥ *v*8.0). All participants who received bedaquiline in the 9-month oral and six-month regimen, received levofloxacin. In this paper we report on secondary analyses of the STREAM Stage 2 trial data to ascertain risk factors for development of QT/QTcF ≥500ms and to validate the ECG monitoring strategy proposed from Stage 1 data.

## Methods

Data from the STREAM Stage 2 trial was used for post-hoc analyses of risk factors for QT/QTcF interval ≥500ms and to assess the accuracy of a monitoring strategy to identify those who developed QT/QTcF interval ≥500ms during follow-up. Participants were included if they were randomised to the 9-month control regimen, the 9-month oral regimen, or a 6-month regimen and received at least 1 dose of trial treatment (safety population). The medications used in each regimen are shown in ***Table 1***. A baseline QT/QTcF ≥450ms was an exclusion criterion in Stage 2.

**Table 1.**
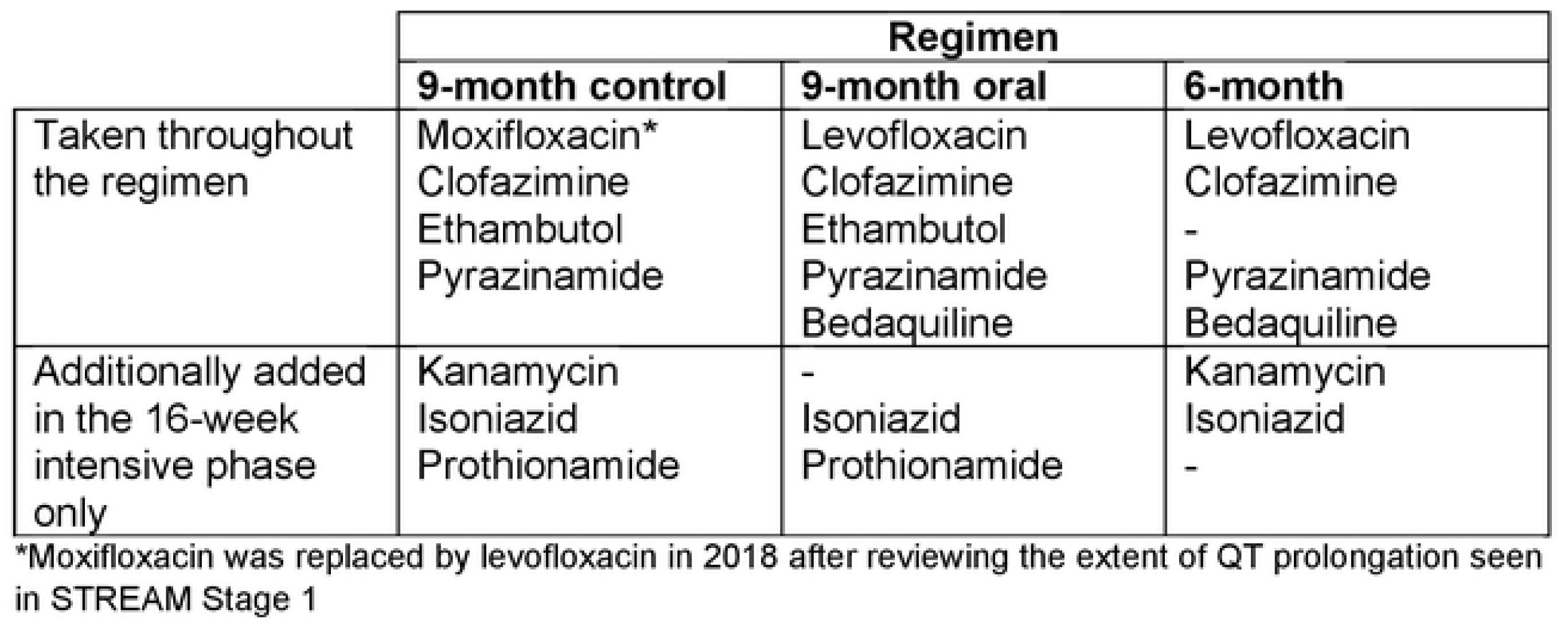
Stream Stage 2 regimen description.

ECGs were recorded at each study visit which was weekly for the first 4 weeks, then 4-weekly until week 52 using standard Eli 250c 12 lead ECG machines at all sites. Five complexes were measured on each lead with lead II being the preferred standard lead for QT measurement. Corrected QT intervals using Fridericia’s formula (QTcF) were calculated from digital ECGs with electronic calipers. Semi-automated, raw waveforms from single leads were used. These were centrally reviewed by a team of cardiologists at IQVIA.

A logistic regression model was fitted to assess the association between a QT/QTcF ≥500ms on any visit with the following baseline variables: age, sex, country, creatinine, QTcF, heart rate (HR), thyroid stimulating hormone (TSH), ALT, grade 1 electrolyte disturbance (potassium ≤ 3.4 mmol/L or magnesium≤ 0.7 mmol/L or calcium≤ 2.10 mmol/L), inclusion of bedaquiline, levofloxacin or moxifloxacin in the assigned regimen, and mean mg/kg clofazimine. An additional analysis was performed to assess whether potassium, magnesium or calcium deficiency during follow up was a risk factor for QT/QTcF interval ≥500ms. End of treatment thyroid function was explored to assess development of hypothyroidism (a known risk factor for QT prolongation).

The evaluation of monitoring strategies was based on the results of analyses from STREAM Stage 1 which found that QTcF thresholds of ≥425ms at 4 hours after the first dose of medication and ≥430ms at week 3 combined, could identify “high-risk” (experienced QT/QTcF ≥500ms during follow-up) and “lower-risk” participants who did not reach 500ms. [7] Analyses assessed the best combination of time points from the different readings within the first month of treatment and tested QTcF thresholds from 410-450ms in 5ms increments. Participants with at least four of six planned ECG measurements up to week 4 of treatment were included. Only QT/QTcF ≥500ms recorded on scheduled monitoring visits were included in the monitoring analyses. QT and QTcF values taken from machine readings were also recorded at each study visit for all participants recruited under version 7.0 (and above) of the protocol; analyses were repeated using the machine readings for this subset of participants.

### Patient Consent Statement

The Union Ethics Advisory Group (EAG) approved (i) the use of deidentified data from the STREAM trials for this post-hoc secondary analysis, and (ii) the waiver of requirement for additional consent from participants (EAG 53/19).

#### Statistical analysis

For the risk factor analyses, continuous variables were described using mean (SD) or median (IQR) while categorical variables were presented as N(%), by prolongation status. Logistic regression models were used to explore factors associated with QT/QTcF ≥500ms; each variable was examined individually then a multivariable model was determined using backwards elimination (exit probability 0.10). Age and country remained in all models as considered a priori to be important. The distribution of continuous variables was assessed graphically. Age, BMI, creatinine, thyroxine, TSH, ALT and AST were log-transformed to achieve normality. An exploratory analysis was performed to assess the effect of bedaquiline on QT/QTcF prolongation after adjusting for other identified risk factors and restricting the analysis to those randomised to a levofloxacin containing regimen to eliminate the possible effects of moxifloxacin.

For the monitoring strategy analyses, participants with missing ECGs were excluded from analysis at that particular time point.

All analyses were performed in Stata 18.0.

## Results

### Predictive risk factors

Of the 556 participants in the safety population, 202 were randomised to the control regimen; 211 the oral regimen and 143 the six-month regimen. Those that reached a QT/QTcF ≥500ms on each regimen is shown in ***Table 2***. The population used for the predictive risk factors analysis is shown in the first row (all visits), and the monitoring strategy analysis in the second row (scheduled visits). The 9-month control regimen contained the largest proportion of participants that reached a QT/QTcF ≥500ms (14 [6.9%], p=0.33). Participants with machine readings available was 219 in total (control n=90; 9-month oral n=98; six-month n=31).

**Table 2.** Number of participants with QT prolongation by regimen.

|  | Regimen |  |  |
| --- | --- | --- | --- |
|  | 9-month control<br>(n=202) | 9-month oral<br>(n=211) | 6-month<br>(n=143) |
| QT/QTcF $\geq 500$ms (all visits) | 14 (6.9%) | 8 (3.8%) | 6 (4.2%) |
| QT/QTcF $\geq 500$ms (scheduled visits) | 11 (5.4%) | 5 (2.4%) | 4 (2.8%) |

***Table 3*** presents baseline characteristics by prolongation event status. Participants with QT/QTcF ≥500ms, had a higher mean [SD] baseline QTcF and higher mean baseline TSH. The highest number of QT/QTcF ≥500ms events were seen in Mongolia (18 [64%]) followed by Moldova (4 [14%]). There was no difference in the distribution of prolongation events by gender; 5% of men and 5% of women reported prolongation ≥500ms. The mean baseline QTcF was 396.3ms (95% CI 394.2, 398.5) in men and 400.8ms (95% CI 397.9, 403.8) in women. The mean change in QTcF from baseline to the maximum seen in follow-up was 53.3ms (95% CI 51.5, 55.1) in the no prolongation group and 98.3ms (95% CI 85.5, 111.0) in participants with QT/QTcF ≥500ms, a 45ms higher maximum change from baseline in the prolongation group compared to the group with no reported prolongation ≥500ms.

**Table 3:** Baseline demographic, medical and laboratory characteristics by QT prolongation.

|  |  | No Prolongation | Prolongation | Total |
| --- | --- | --- | --- | --- |
| Country | Ethiopia | 61 (100%) | 0 | 61 (100%) |
|  | South Africa | 84 (98%) | 2 (2%) | 86 (100%) |
|  | Mongolia | 101 (85%) | 18 (15%) | 119 (100%) |
|  | Moldova | 55 (93%) | 4 (7%) | 59 (100%) |
|  | Georgia | 31 (97%) | 1 (3%) | 32 (100%) |
|  | Uganda | 55 (98%) | 1 (2%) | 56 (100%) |
|  | India | 141 (99%) | 2 (1%) | 143 (100%) |
| Age - years |  | 34.8 ± 11.3 | 35.4 ± 10.9 | 34.8 ± 11.3 |
| Sex | Male | 320 (95%) | 17 (5%) | 337 (100%) |
|  | Female | 208 (95%) | 11 (5%) | 219 (100%) |
| BMI kg/m<sup>2</sup> |  | 19.8 ± 4.0 | 19.7 ± 3.0 | 19.8 ± 3.9 |
| HIV status | Negative | 439 (94%) | 26 (6%) | 465 (100%) |
|  | Positive | 89 (98%) | 2 (2%) | 91 (100%) |
| ART status | No | 469 (95%) | 26 (5%) | 495 (100%) |
|  | Yes | 59 (97%) | 2 (3%) | 61 (100%) |
| Diabetes | No | 481 (95%) | 27 (5%) | 508 (100%) |
|  | Yes | 47 (98%) | 1 (2%) | 48 (100%) |
| QTcF - ms |  | 397.0 ± 20.7 | 419.1 ± 18.3 | 398.1 ± 21.2 |
| HR - bpm |  | 95.4 ± 16.8 | 84.0 ± 16.4 | 94.8 ± 16.9 |
| Creatinine – umol/L |  | 63.8 ± 17.2 | 66.9 ± 14.2 | 63.9 ± 17.0 |
| Potassium – mmol/L |  | 4.4 ± 0.5 | 4.4 ± 0.4 | 4.4 ± 0.5 |
| Calcium – mmol/L |  | 2.3 ± 0.1 | 2.3 ± 0.1 | 2.3 ± 0.1 |
| Magnesium – mmol/L |  | 0.8 ± 0.1 | 0.9 ± 0.1 | 0.8 ± 0.1 |
| TSH – mIU/L |  | 1.9 ± 1.4 | 2.9 ± 2.2 | 2.0 ± 1.4 |
| Thyroxine – nmol/L |  | 109.4 ± 28.3 | 101.4 ± 22.4 | 109.0 ± 28.0 |
| ALT – u/L |  | 26.7 ± 26.3 | 23.4 ± 16.8 | 26.5 ± 25.9 |
| AST – u/L |  | 29.5 ± 20.8 | 27.2 ± 14.5 | 29.4 ± 20.5 |

Data are n (%) or mean ± SD; BMI – body mass index, HIV = human immunodeficiency virus, ART = antiretroviral therapy, HR = heart rate, QTcF = corrected QT interval Fridericia's formula, TSH = thyroid stimulating hormone, ALT = alanine transaminase, AST = aspartate transaminase, SD = standard deviation.

***Table 4*** presents the results of the univariable and multivariable regressions. The final model included country, baseline QTcF, baseline TSH and the allocated fluoroquinolone (levofloxacin or moxifloxacin). With regards to the country effect, the models show the odds of QT/QTcF ≥500ms were not significantly different between Mongolia, Moldova and Georgia, but they are significantly higher in Mongolia than in India and the African countries. We also included adjustment for age but it was not statistically significant. There was some association between QT/QTcF ≥500ms in those who received moxifloxacin as opposed to levofloxacin (OR 2.49; 95% CI 0.92 to 6.71, p = 0.07).

**Table 4.**
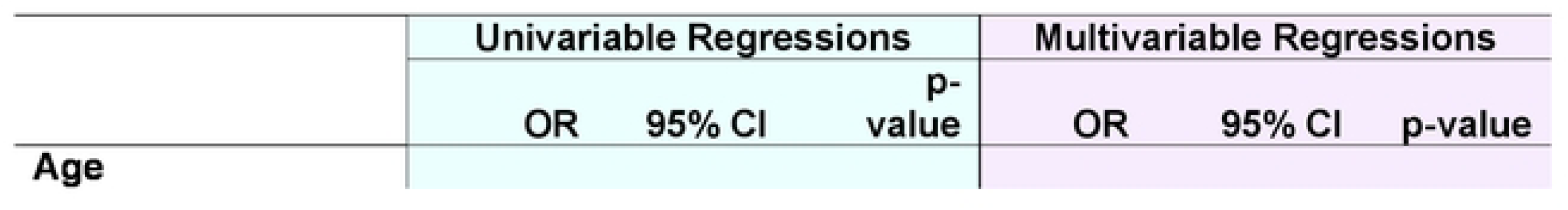
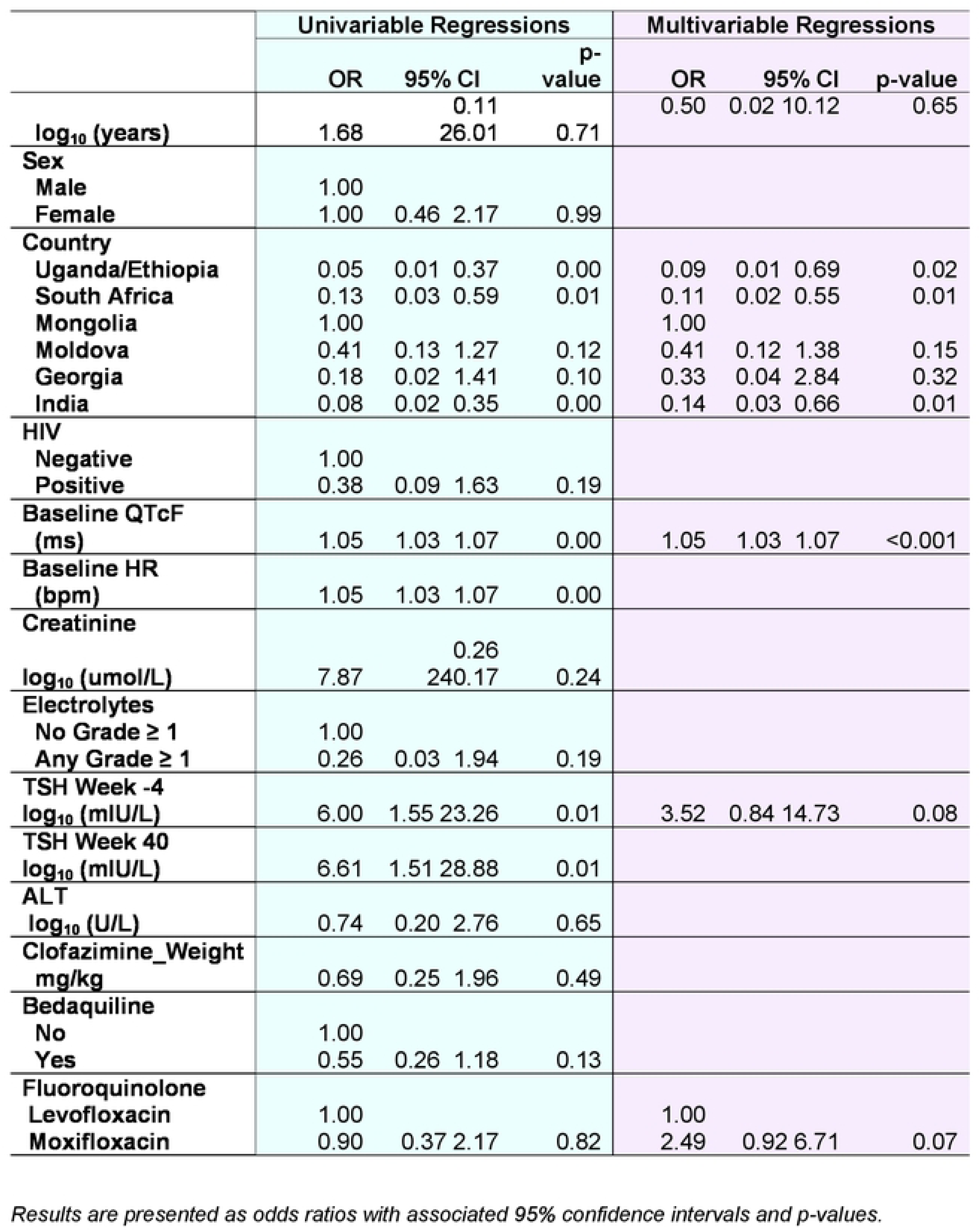
Univariable and Multivariable regressions.

There was some association between a higher baseline TSH level and increased odds of QT/QTcF ≥500ms (OR 3.52; 95% CI 0.84 to 14.73, p = 0.08). This association persisted at the end of treatment. TSH change between baseline and the end of treatment was similar in the population that experienced a QT/QTcF ≥500ms and those who did not (Mean (SD)_QT_ = 0.37 (1.86) vs Mean (SD)_No_QT_=0.36 (1.30), t-test p-value = 0.98) ***(Table S1)*.**

Univariable analysis showed some suggestion that bedaquiline was associated with reduced odds of QT/QTcF ≥500ms [OR 0.55; 95% CI 0.26 to 1.18, p = 0.13 (***Table 4***)], however as the number of events was small this could have influenced the OR. After adjusting for other associated factors in the final multivariable model and restricting the analysis to those allocated levofloxacin, although the odds of QT/QTcF ≥500ms were increased for those taking bedaquiline there was no statistically significant association (OR = 1.19; 95% CI 0.36 to 3.95, p=0.77).

A further analysis investigated the relationship between DAIDS Grade 1 electrolyte deficiencies during trial follow-up and development of a QT/QTcF ≥500ms (***Table S2***). At baseline, only one participant (1/28, 4%) in the QT/QTcF ≥500ms group had a Grade ≥1 electrolyte abnormality compared with 66/528, (13%); p=0.16, in the no QT/QTcF ≥500ms group. Sixteen participants (16/28, 57%), who experienced a QT/QTcF ≥500ms event, experienced at least one Grade ≥1 electrolyte abnormality during follow-up compared with 344/528 (65%); p=0.39, in the group who didn’t reach a QT/QTcF ≥500ms.

The relationship between the timing of an electrolyte abnormality and a QT/QTcF ≥500ms event was also explored (***Figure S1***). Of 28 patients, nine (32%) had a Grade 1 electrolyte deficiency prior to a QT/QTcF ≥500ms event, three (11%) in the same ECG window, four (14%) after their QT event and twelve patients (43%) had no electrolyte abnormality (Grade ≥1) but still developed QT/QTcF ≥500ms. Roughly half (54%) of participants had a blood test on the day of their QT/QTcF ≥500ms event.

### Monitoring strategy

Of the 556 participants in the monitoring strategy analysis, 11 participants (control regimen = 5; oral regimen = 6) had fewer than four ECG recordings in the first 4 weeks and were excluded. None had a QT/QTcF ≥500ms during the trial.

#### Validation of STREAM Stage 1 monitoring strategy

Combining the two thresholds as in the Stage 1 ECG monitoring strategy, i.e. a QTcF threshold of 425ms at 4-hours and 430ms at week 3 (restricted to those <425ms at 4-hours) gave a sensitivity of 100%; specificity of 62%; PPV 13% and NPV 100%.

A QTcF cut-off threshold of 425ms at 4-hours identified all 11 participants who reached a QT/QTcF ≥500ms during follow-up with 70% (130/186) of participants who never reached a QT/QTcF ≥500ms being below this threshold (***Table 5***). This gave a sensitivity of 100%; specificity of 70%; positive predictive value (PPV) of 16% and a negative predictive value (NPV) of 100%.

**Table 5.** Performance of the STREAM Stage 1 ECG monitoring strategy in the control regimen for STREAM Stage 2 for predicting QT prolongation at up to week 52.

|  | No QT<br>prolongation<br>N (%) | QT prolongation<br>N (%) | Overall<br>N (%) | P-value |
| --- | --- | --- | --- | --- |
| 4-hour ECG reading |  |  |  |  |
| QTcF <425ms | 130 (70) | 0 (0) | 130 (66) | <0.001 |
| QTcF ≥425ms | 56 (30) | 11 (100) | 67 (34) |  |
| Week 3 ECG reading |  |  |  |  |
| QTcF <430ms | 138 (75) | 1 (9) | 139 (71) | <0.001 |
| QTcF ≥430ms | 47 (25) | 10 (91) | 57 (29) |  |
| Week 3 analysis restricted to participants <425ms at 4-hour reading |  |  |  |  |
| QTcF <430ms | 114 (88) | - | 114 (88) |  |
| QTcF ≥430ms | 15 (12) | - | 15 (12) |  |

A QTcF cut-off threshold of 430ms at week 3 (with no adjustment for the 4-hour result), was met by 91% (10/11) of participants who reached a QT/QTcF ≥500ms during follow-up, with 75% (138/185) in the no prolongation group being below this threshold. This gave a sensitivity of 91%; specificity of 75%; PPV of 18% and a NPV of 99%.

#### Performance of STREAM Stage 1 strategy in the oral regimen

The same strategy was applied to the population assigned the 9-month oral regimen (***Table 6***). This gave a sensitivity of 100%; specificity of 59%; PPV 6% and NPV 100%.

**Table 6.** Performance of the STREAM Stage 1 ECG monitoring strategy in the oral regimen for STREAM Stage 2 for predicting QT prolongation at up to week 52.

|  | No QT prolongation<br>N (%) | QT prolongation<br>N (%) | Overall<br>N (%) | P-value |
| --- | --- | --- | --- | --- |
| 4-hour ECG reading |  |  |  |  |
| QTcF <425ms | 162 (81) | 1 (20) | 163 (80) | 0.007 |
| QTcF ≥425ms | 38 (19) | 4 (80) | 42 (20) |  |
| Week 3 ECG reading |  |  |  |  |
| QTcF <430ms | 132 (67) | 1 (20) | 133 (66) | 0.049 |
| QTcF ≥430ms | 66 (33) | 4 (80) | 70 (34) |  |
| Week 3 ECG reading restricted to participants <425ms at 4-hour reading |  |  |  |  |
| QTcF <430ms | 118 (73) | - | 118 (73) | 0.272 |
| QTcF ≥430ms | 43 (27) | 1 (100) | 44 (27) |  |

At the 4-hour time point, Eighty-one percent (162/200) of the low-risk group were below the 425ms threshold. All but one of the five participants who reached a QT/QTcF ≥500ms during the trial were above the 425ms threshold. This gave a sensitivity of 80%; specificity 81%; PPV 10% and NPV 99%.

A QTcF cut-off threshold of 430ms at week 3 (with no adjustment for the 4-hour result), was met by 80% (4/5) participants who reached a QT/QTcF ≥500ms during follow-up, with 67% (132/198) in the no prolongation group being below this threshold. This gave a sensitivity of 80%; specificity of 67%; PPV of 6% and a NPV of 99%.

#### Exploration of alternative monitoring strategy in the oral regimen

Analysis of a single time point at week 4 using a QTcF cut-off threshold of 435ms is shown in ***Table 7***. Monitoring could have been reduced in 69% (137/198) of the lower risk group. All five of the participants who developed QT/QTcF ≥500ms were above the 435ms cut-off.

**Table 7.** Performance of the alternative monitoring strategy in the oral regimen.

|  | No QT prolongation<br>N (%) | QT prolongation<br>N (%) | Overall<br>N (%) | P-value |
| --- | --- | --- | --- | --- |
| Week 4 ECG reading |  |  |  |  |
| QTcF <435ms | 137 (69) | 0 (0) | 137 (67) | 0.003 |
| QTcF ≥435ms | 61 (31) | 5 (100) | 66 (33) |  |

#### Performance of STREAM Stage 1 strategy in the six-month regimen

The same analysis for the population assigned the 6-month regimen is shown in the appendix (***Table S3***). Following the STREAM Stage 1 strategy, a sensitivity 75%; specificity 63%; PPV 5% and NPV 99% was achieved.

#### Performance of STREAM Stage 1 strategy using machine readings

The same strategy was also applied to the machine readings that were available (***Tables S4-S6***). In summary, the strategy performed less well.

## Discussion

### Risk factors contributing to QT prolongation

Baseline QTcF and country were significant risk factors for developing a QT/QTcF ≥500ms in the STREAM Stage 2 population. This supports findings from STREAM Stage 1 analyses. [3] A baseline QTcF ≥400ms was associated with increased odds of reaching a QT/QTcF ≥500ms. It seems plausible that starting with a higher QTcF before taking medications known to prolong the QT interval would increase the chance of reaching 500ms. Mongolia was again found to have a higher proportion of participants who developed a QT/QTcF ≥500ms; the explanation for this is unknown but seems likely related to genetic or environmental factors.[3] Participants recruited close to Ulaanbaatar for the GENDISCAN study had genetic anomalies in the chromosomal regions encoding the potassium channel genes KCNH2 and KCND2 which control the QT interval. [9] Genetic anomalies in the potassium channel gene KCNQ1 have also been demonstrated in other Mongolian populations. [10] It is possible that the addition of environmental risk factors such as extreme cold and contaminated drinking water could potentiate any genetic abnormality in the above genes for this geographic region. [10,11] A higher risk of QT prolongation has also been demonstrated in another central Asian population in the TB-PRACTECAL trial and in an east Asian population in Taiwan. [12,13]

Analysis of drug effects on the QT interval was complicated by different regimens having different combinations of drugs and all regimens including some core drugs, such as clofazimine, so it was difficult to disentangle the impact of individual agents. There was a suggestion of increased odds of developing a QT/QTcF ≥500ms in participants randomised to moxifloxacin compared with those assigned levofloxacin. Though QT/QTcF ≥500ms is associated with all fluoroquinolones, moxifloxacin has been shown to carry the greatest risk, particularly at higher doses [14] and this effect is increased by the co-administration of clofazimine. [15]

Assessment of the bedaquiline effect on QT/QTcF ≥500ms was not straightforward as the oral and six-month regimens also contained levofloxacin and clofazimine, with concomitant use of the later known to increase the QT interval compared to bedaquiline alone. [16] Surprisingly, univariable analysis suggested reduced odds of QT/QTcF ≥500ms with bedaquiline but this was likely driven by the higher risk associated with moxifloxacin in the control regimen, which didn’t contain bedaquiline. When the analysis was restricted to those allocated a levofloxacin-containing regimen i.e. control regimen participants on levofloxacin and all those on oral and six-month regimens, there were slightly increased odds of QT/QTcF ≥500ms in bedaquiline containing regimens, although not statistically significant.

Hypothyroidism is increasingly recognised in MDR-TB populations. [17–19]. We found a higher baseline TSH was associated with six times the odds of QT/QTcF ≥500ms in the univariable model, although not significant in the multi variable model. Hypothyroidism is a known risk factor for QT prolongation. [19,20] The control and oral regimens in STREAM Stage 2 include protionamide, which is associated with development of hypothyroidism possibly due to its structural similarity to carbimazole. [18]

### Validation of ECG monitoring strategy

The monitoring strategy developed from Stage 1 data performed well in the control regimen. In contrast to Stage 1, an ECG at 4 hours after the first dose of treatment in this data set identified all the high-risk participants; if validated, it could potentially allow reduced monitoring in 70% of participants. The second time point did not identify more of the high-risk group, but reduced the false positivity rate for the low-risk group. Interestingly, though a single time point identified all the high-risk participants in this dataset, it was not consistent with the Stage 1 analyses and would need to be validated in a larger dataset to be confident the finding was reproducible.

Since STREAM Stage 2 started, there has been a move away from the use of injectables. [4,5,21] Assessment of the monitoring strategy in an all-oral bedaquiline containing regimen was therefore important. Our analysis showed the Stage 1 strategy performed very well in the oral regimen. This was unexpected good news, given the unusual pharmacokinetics of bedaquiline whereby a steady state in plasma is not reached until week 2 and beyond. [22]

We found an alternative strategy also performed well in the oral regimen. A single time-point one month into treatment with a cut-off of 430ms was identified. If validated, this may be easier to implement in programmatic settings than the 4-hour and week 3 time points.

The monitoring strategy did not perform as well on the machine readings with false negatives observed across the regimens. As already described, there were fewer ECGs available (less than 50%). It is possible that the machine readings of the QT interval were less accurate than the cardiologist read calculations and is a known issue with ECGs. [23–25] Two of the regimens also contained drugs different to those used in STREAM Stage 1 which again may have affected the performance. The ECG machines used in Stage 1 (GE MAC 800) and Stage 2 (ELI 250c) differed, which could also explain the why the results for the machine readings were not reproducible.[24,26]

Strengths

1. In addition to the baseline risk factors described in Stage 1, follow-up bloods for electrolyte deficiencies were assessed.
2. Baseline and end of treatment thyroid function were assessed.
3. The strategy developed from Stage 1 data was applied to a different population in a wider geographic area who took the same regimen and still performed well.
4. The monitoring strategy was assessed in patients taking different regimens to those in Stage 1 including two bedaquiline containing regimens.

Limitations

1. This analysis did not include pharmacokinetic data so is unable to explore the relationship between plasma concentrations of moxifloxacin, levofloxacin, clofazimine and bedaquiline and the effect on the QT interval.
2. Analyses of the monitoring strategy for Stage 2 data relied on a central cardiologist who manually calculated the QT and QTcF interval as opposed to the original strategy developed from STREAM Stage 1 data that used machine calculations of the QT and QTcF interval.

The implications from this work are that we have identified significant baseline risk factors that increase patients’ risk of QT/QTcF ≥500ms, a risk factor for potentially life threatening arrythmias. Our proposed monitoring strategy has been shown to safely reduce the number of ECG visits for patients without missing those who need closer monitoring. The 9-month oral regimen is now recommended by WHO for the treatment of rifampicin-resistant TB so these results are of importance in the planning of monitoring strategies in programmatic settings.

## Data Availability

The authors agree to make data fully available when requested.

## Acknowledgements

We thank all the participants, community action board members, and collaborators without whom the STREAM study would not have been possible.

## References

[1] World Health Organization. WHO consolidated guidelines on tuberculosis. Module 4: treatment – drug-resistant tuberculosis treatment. Online annexes. 2020.

[2] Nunn AJ, Phillips PPJ, Meredith SK, Chiang C-Y, Conradie F, Dalai D, et al. A Trial of a Shorter Regimen for Rifampin-Resistant Tuberculosis. NEJM 2019:1201–13. 10.1056/nejmoa1811867.

[3] Hughes G, Bern H, Chiang CY, Goodall RL, Nunn AJ, Rusen ID, et al. QT prolongation in the STREAM Stage 1 Trial. Int J Tuberc Lung Dis 2022;26:334–40. 10.5588/ijtld.21.0403.

[4] Nyang’wa B-T, Berry C, Kazounis E, Motta I, Parpieva N, Tigay Z, et al. A 24-Week, All-Oral Regimen for Rifampin-Resistant Tuberculosis. NEJM 2022;387:2331–43. 10.1056/nejmoa2117166.

[5] Conradie F, Bagdasaryan TR, Borisov S, Howell P, Mikiashvili L, Ngubane N, et al. Bedaquiline– Pretomanid–Linezolid Regimens for Drug-Resistant Tuberculosis. NEJM 2022;387:810–23. 10.1056/nejmoa2119430.

[6] Paton NI, Cousins C, Suresh C, Burhan E, Chew KL, Dalay VB, et al. Treatment Strategy for Rifampin-Susceptible Tuberculosis. NEJM 2023;388:873–87. 10.1056/nejmoa2212537.

[7] Hughes G, Bern H, Chiang CY, Goodall RL, Nunn AJ, Rusen ID, et al. ECG monitoring in STREAM Stage 1: can we identify those at increased risk of QT prolongation? Int J Tuberc Lung Dis 2022;26:1065–70. 10.5588/ijtld.22.0063.

[8] Goodall RL, Meredith SK, Nunn AJ, Bayissa A, Bhatnagar AK, Bronson G, et al. Evaluation of two short standardised regimens for the treatment of rifampicin-resistant tuberculosis (STREAM stage 2): an open-label, multicentre, randomised, non-inferiority trial. The Lancet 2022;400:1858–68. 10.1016/S0140-6736(22)02078-5.

[9] Im SW, Lee MK, Lee HJ, Oh S Il, Kim HL, Sung J, et al. Analysis of genetic and non-genetic factors that affect the QTc interval in a Mongolian population: The GENDISCAN study. Exp Mol Med 2009;41:841–8. 10.3858/emm.2009.41.11.090.

[10] Odgerel Z. Genetic variants in potassium channels are associated with type 2 diabetes in Mongolian population. J Diabetes 2012;4:238–42. 10.1038/jid.2014.371.

[11] Mumford JL, Wu K, Xia Y, Kwok R, Yang Z, Foster J, et al. Chronic arsenic exposure and cardiac repolarization abnormalities with QT interval prolongation in a population-based study. Environ Health Perspect 2007;115:690–4. 10.1289/ehp.9686.

[12] Motta I, Cusinato M, Ludman AJ, Lachenal N, Dodd M, Soe M, et al. How much should we still worry about QTc prolongation in rifampicin-resistant tuberculosis? ECG findings from TB-PRACTECAL clinical trial. Antimicrob Agents Chemother 2024. 10.1128/aac.00536-24.

[13] Lin CJ, Chen JH, Chien ST, Huang YW, Lin C Bin, Lee JJ, et al. Clofazimine and QT prolongation in the treatment of rifampicin-resistant tuberculosis: Findings of aDSM in Taiwan. J. Microbiol. Immunol. Infect 2024. 10.1016/j.jmii.2024.08.002.

[14] Falagas ME, Rafailidis PI, Rosmarakis ES. Arrhythmias associated with fluoroquinolone therapy. Int J Antimicrob Agents 2007;29:374–9. 10.1016/j.ijantimicag.2006.11.011.

[15] Radtke KK, Hesseling AC, Winckler JL, Draper HR, Solans BP, Thee S, et al. Moxifloxacin Pharmacokinetics, Cardiac Safety, and Dosing for the Treatment of Rifampicin-Resistant Tuberculosis in Children. Clin. Infect. Dis 2022;74:1372–81. 10.1093/cid/ciab641.

[16] Brust JCM, Gandhi NR, Wasserman S, Maartens G, Omar S V., Ismail NA, et al. Effectiveness and Cardiac Safety of Bedaquiline-Based Therapy for Drug-Resistant Tuberculosis: A Prospective Cohort Study. Clin. Infect. Dis 2021;73:2083–92. 10.1093/cid/ciab335.

[17] Modongo C, Zetola NM. Prevalence of hypothyroidism among MDR-TB patients in Botswana. Int J Tuberc Lung Dis 2012;16:1561–2. 10.5588/ijtld.12.0403.

[18] Cheung YMM, Van K, Lan L, Barmanray R, Qian SY, Shi WY, et al. Hypothyroidism associated with therapy for multi-drug-resistant tuberculosis in Australia. Intern Med J 2019;49:364–72. 10.1111/imj.14085.

[19] Hee Kweon K, Hyun Park B, Gu Cho C. Address for correspondence The Effects of L-thyroxine Treatment on QT Dispersion in Primary Hypothyroidism. J Korean Med Sci 2007;22:114–20.

[20] Altun A Agobkmog. The relationship between ventricular repolarization and thyroid stimulating hormone. Ann Noninvasive Electrocardiogr 1998;3.

[21] WHO. Rapid communication: Key changes to the treatment of drug-resistant tuberculosis. 2022.

[22] van Heeswijk RPG, Dannemann B, Hoetelmans RMW. Bedaquiline: A review of human pharmacokinetics and drug-drug interactions. JAC 2014;69:2310–8. 10.1093/jac/dku171.

[23] Barbey JT, Connolly M, Beaty B, Krantz MJ. Man versus Machine: Comparison of Automated and Manual Methodologies for Measuring the QTc Interval: A Prospective Study. Ann Noninvasive Electrocardiogr 2016;21:82–90. 10.1111/anec.12277.

[24] Darpo B, Agin M, Kazierad DJ, Layton G, Muirhead G, Gray P, et al. Man versus machine: Is there an optimal method for QT measurements in thorough QT studies? J Clin Pharmacol 2006;46:598–612. 10.1177/0091270006286900.

[25] Savelieva I, Gang Y, Guo XH, Hnatkova K, Malik M. Agreement and reproducibility of automatic versus manual measurement of QT interval and QT dispersion. AJC 1998;81:471–7. 10.1016/S0002-9149(97)00927-2.

[26] Sano M, Aizawa Y, Katsumata Y, Nishiyama N, Takatsuki S, Kamitsuji S, et al. Evaluation of differences in automated QT/QTc measurements between Fukuda Denshi and Nihon Koden systems. PLoS One 2014;9. 10.1371/journal.pone.0106947.

